# Adenoviral Vectors Overcome Immunosuppression Via Antigen Persistence and Metabolic Reprogramming

**DOI:** 10.64898/2026.03.05.26347734

**Authors:** Shixiong Li, Shihao Shang, Kepu Liu, Chengshu Xie, Qiong Zi, Zumrat Nur, Danni Feng, Maoxin Lv, Zelong Sun, Di Wei, Yuhao Yan, Jianguo Deng, Jianlin Yuan, Zhang-Yong Hong, Yingjie Bian, Zheng Zhu, Jingyou Yu

**Affiliations:** Guangzhou National Laboratory, Bio-Island, Guangzhou, Guangdong 510005, China; Department of Urology, Xijing Hospital, Fourth Military Medical University,710032, Xi’an, China; College of Life Sciences, Nankai University, Tianjin 300071, China; State Key Laboratory of Respiratory Disease, The First Affiliated Hospital of Guangzhou Medical University, Guangzhou 510182, China

**Keywords:** Adenoviral vectors, immunosuppression, pre-existing immunity, antibody response, metabolic reprogramming, kidney transplant recipients, vaccine platform comparison

## Abstract

Vaccination frequently elicits suboptimal immunogenicity in organ transplant recipients, particularly those on long-term immunosuppressive therapy, highlighting the need for improved understanding of immunosuppression mechanisms and optimized vaccination strategies. This study enrolled a cohort of 132 individuals and observed significantly lower antibody levels in kidney transplant recipients (KTRs) compared to non-transplant controls (non-KTRs). Antibody levels were inversely associated with both the dosage and duration of immunosuppressive therapy. Complementary small animal studies demonstrated that immunosuppressive treatment dosage-dependently and reversibly impaired antibody production, primarily by depleting immune cells, notably B cells. A single shot of adenoviral vector-based vaccines demonstrated enhanced immunogenicity relative to two shots of alum-adjuvanted protein vaccines, inducing potent neutralizing antibodies (NAbs) and a Th1-biased T-cell response even under continuous immunosuppression. The enhanced response was driven by reduced interference from pre-existing antibodies, sustained transgene expression, and the reprogramming of lipid metabolism to activate T and B cells. Our findings advocate for tailored vaccination strategies, positioning adenoviral vectors as a candidate modality for this vulnerable population.

## INTRODUCTION

Vaccination remains a primary strategy for mitigating severe disease and mortality associated with infectious diseases. Despite significant advancements in vaccine platform development, suboptimal immunogenicity, particularly among immunocompromised individuals, persists as a critical challenge. Patients undergoing immunosuppressive therapy, such as solid organ transplant recipients, face substantially elevated risks of severe complications following SARS-CoV-2 infection ^1^. Prolonged SARS-CoV-2 replication in immunocompromised individuals has raised concerns about the adequacy of immunologic control in these populations and, more critically, facilitated the emergence of new variants, as likely seen with Omicron ^2^. Therefore, assessing the immune responses elicited by COVID-19 vaccines in these vulnerable cohorts is not only necessary for individual protection but also essential to guiding public health policies and vaccination strategies.

Kidney transplant recipients constitute a substantial proportion of all solid organ transplant recipients and represent a key immunocompromised cohort, necessitating lifelong immunosuppressive therapy to maintain graft function and prevent rejection ^3^. This population is notably more susceptible to infections and typically experiences diminished vaccine responses compared to healthy individuals ^4–6^. We and others have recently revealed that solid organ transplant recipients, including kidney transplant recipients, exhibit reduced serological responses following vaccination against COVID-19 ^7–14^. Alarmingly, after two doses of mRNA COVID-19 vaccines, only 38% to 54% of kidney and liver transplant recipients develop detectable SARS-CoV-2 antibodies ^15–17^. While additional mRNA vaccine doses improve responses for some, many patients retain suboptimal immunity, directly attributable to ongoing immunosuppressive therapy ^8,18–23^. Key determinants of this attenuated immunogenicity include the types and dosages of immunosuppressive drugs being administered ^5^, the timing of the vaccine relative to transplant surgery or dialysis ^24^, and inherent individual variations in immune competence ^25–27^.

Although blunted antibody responses to mRNA COVID-19 vaccines are well-documented in transplant recipients under intensive immunosuppression, a critical gap exists regarding inactivated or protein-based vaccines, which are extensively and globally deployed ^28^. The efficacy of these platforms in immunosuppressed populations remains inadequately characterized. Complicating this, the nature and potency of hybrid immunity (resulting from vaccination plus natural infection) in immunosuppressed individuals, and how it might be influenced by the vaccine platform, remain poorly defined, warranting urgent and comprehensive investigation.

Emerging evidence also suggests that specific vaccine platforms confer unique immunological advantages in immunocompromised hosts. Adenoviral vectored vaccines, for instance, demonstrate compelling features associated with potentially superior immunogenicity compared to traditional inactivated or protein-based vaccines ^29,30^. Adenoviral vectored vaccines, such as those based on Ad5, are known to elicit robust and balanced humoral and cellular immune responses in immunocompetent individuals, partly due to their inherent adjuvant properties and efficient transduction of host cells. These advantages likely stem from intrinsic adjuvanticity, which potently activates innate immune pathways; highly efficient cell entry facilitating endogenous antigen production; and comprehensive antigen presentation via both MHC class I and II molecules. This enables robust stimulation of both cytotoxic CD8+ T-cells and helper CD4+ T-cells alongside humoral responses, culminating in a balanced Th1/Th2 immune profile. This coordinated cellular and humoral immunity is frequently linked to enhanced overall immunogenicity and potentially more durable protection ^31^, attributes critically needed where traditional vaccines often fail. While other platforms like mRNA vaccines also elicit balanced responses, adenoviral vectors leverage a distinct mechanism of action based in viral delivery and inherent adjuvant properties. Clarifying the precise mechanisms underlying the relative efficacy of these platforms in vulnerable populations will be paramount for designing next-generation vaccines and optimizing immunization strategies.

This study investigates antibody responses elicited by inactivated COVID-19 vaccines within two distinct cohorts: kidney transplant recipients (KTRs) and non-transplant controls (non-KTRs). We specifically examine the impact of diverse immunosuppressive regimens, dialysis status, and time elapsed since transplantation on vaccine immunogenicity. We further explore strategies aimed at augmenting immune responses under conditions of immunosuppression. Our findings confirm the detrimental impact of triple-drug immunosuppressive regimens on antibody titers, reinforcing prior observations on factors influencing vaccine response. Crucially, we demonstrate that adenoviral vectored vaccines induce a uniquely advantageous immune profile in this immunosuppressed population.

## RESULTS

### Kidney transplant recipients (KTRs) exhibit significantly reduced antibody levels compared to immunocompetent controls

To assess the antibody responses in kidney transplant recipients (KTRs) and immunocompetent Non-KTR, we conducted a cohort study in 132 individuals ≥18 years of age who received three inactivated vaccines (Sinopharm or CoronaVac) or unvaccinated from May 7, 2021 through December 25, 2022 (Fig. 1A and Table 1). Immune responses were assessed 25.8 months (interquartile range: 22.9-34.6) after the last dose. 33 out of 40 KTRs underwent SARS-CoV-2 breakthrough infection (82.5%), while 87 out of 92 non-KTRs reported a symptomatic infection (94.6%). The reported symptomatic infections have been confirmed by serological ELISA tests against SARS-CoV-2 N protein (Fig. S1A). Notably, KTR cohorts routinely take dialysis two years before take the kidney transplant and immunosuppressant simultaneously (Table 2), which maintained thereafter regardless of vaccination or breakthrough infection (Fig. 1A).

**Figure 1.**
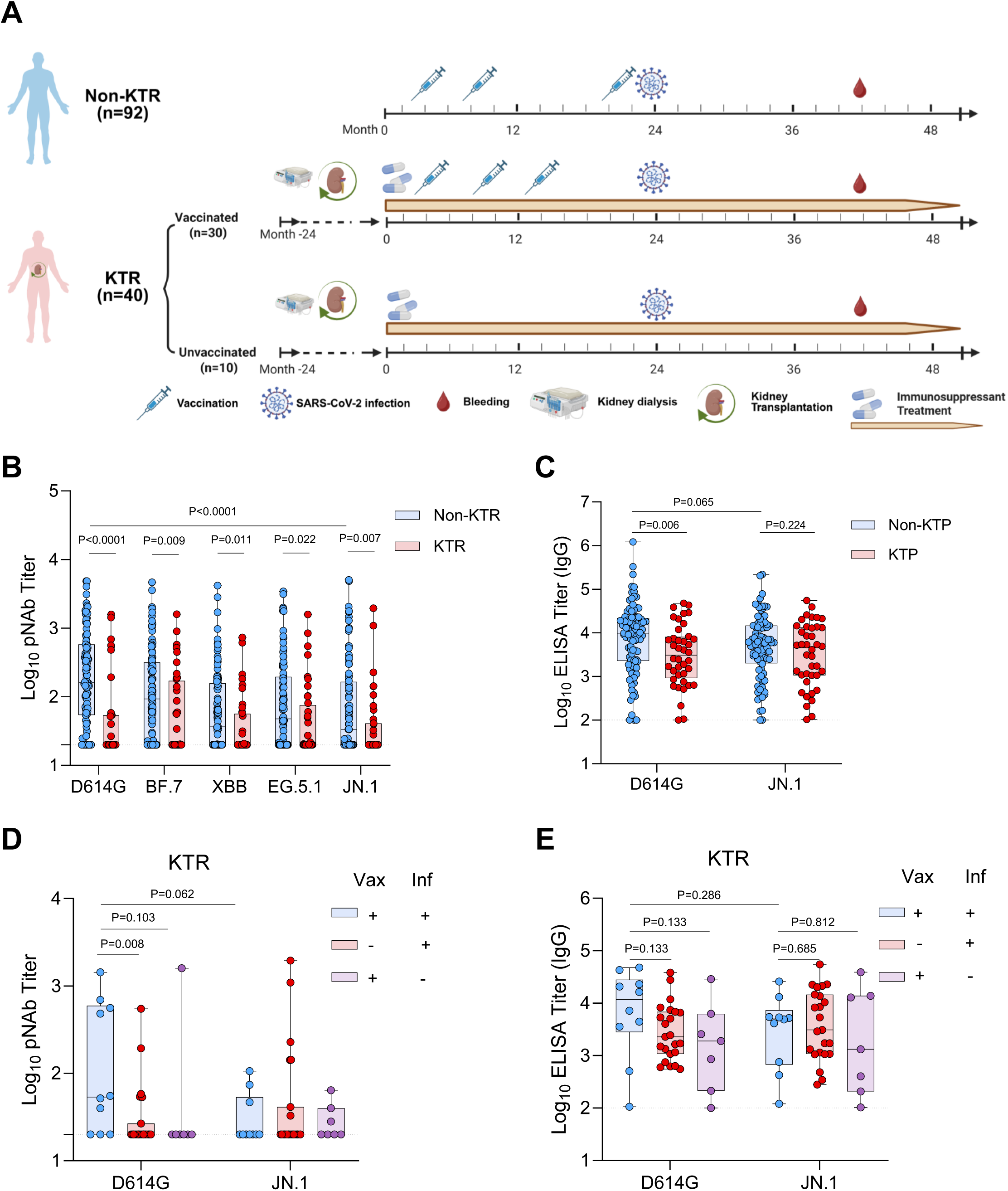
Kidney transplant recipients (KTRs) exhibit impaired humoral responses to SARS-CoV-2 vaccination. (A) Schematic timeline of the clinical cohort study. Day 0 represents the date of the first SARS-CoV-2 vaccination. Sample sizes (N) for kidney transplant recipients (KTRs) and non-transplant controls (Non-KTRs) who received three doses of inactivated vaccine are indicated. KTRs underwent dialysis prior to transplantation and initiated immunosuppressant (IS) therapy at transplantation. (B) Neutralizing antibody (NAb) titers against various SARS-CoV-2 variants, including ancestral strain D614G and variants BF.7, XBB, EG.5.1, and JN.1, as determined by pseudovirus neutralization assay. (C) Serum IgG titers against the receptor-binding domain (RBD) of D614G and JN.1, measured by enzyme-linked immunosorbent assay (ELISA). Serum neutralizing antibody (D) and RBD-specific IgG (E) titers in KTRs, stratified by vaccination and SARS-CoV-2 infection history. Groups: Vaccinated & Infected (Vax+ Inf+, n=10), Unvaccinated & Infected (Vax-Inf+, n=23), Unvaccinated & Uninfected (Vax-Inf-, n=7). For (B-E), data are presented as median ± interquartile range (IQR). Statistical significance was determined by two-sided Mann-Whitney U tests. The dashed line indicates the lower limit of quantification (LLOQ: 20 for NAb, 100 for ELISA).

Neutralizing antibody (NAb) responses were detected in 83.7% of immunocompetent controls. However, only 65.0% of individuals in the KTR group had detectable NAb titers against the ancestral strain D614G (Fig. 1B). The median NAb titer was 161.8 in non-KTRs, 20 in the KTR group (p<0.0001, two-sided Mann-Whitney U tests) (Fig. 1B). Neutralizing antibody responses were detected against BF.7, XBB, EG.5.1, and JN.1, four variants that once circulated in Mainland China, in the non-KTR group, with median NAb titers of 93.2, 36.5, 47.8, 33.5, respectively. NAb titers against variants were all <20 in the KTR group (Fig. 1B and S1B). Median RBD binding titers against D614 and JN.1 in the KTR group were also markedly reduced (Fig. 1C). Notably, antigenic distances, calculated from neutralization titers, showed no statistically significant difference between D614G and the four variants after the removal of confounding non-neutralizers (Fig. S1C-D), indicating a qualitative similarity in antibody responses between KTR and non-KTR individuals. The results thus confirmed results from our group and others that kidney transplants receivers who took immunosuppressants induced a markedly lower level of serologic antibody responses than the immunocompetent counterparts.

### KTR individuals experiencing breakthrough infection demonstrated higher serum antibody levels

Breakthrough infection has reported to be able boost immunity against invading viral pathogens^32^. We aimed to assess whether this still holds true in immunosuppressed individuals. In KTR, individuals who were unvaccinated or uninfected showed a NAb titer below of lower limit of detection (<20) for both D614G and JN.1 variants (Fig. 1D). Infection-only failed to significantly boost NAb titer, while vaccination plus breakthrough infection did elevate median NAb to 53.5 (Fig. 1D). Similar trend could be extended to RBD-specific binding antibody responses, with median titers of D614G and JN.1 was 1,899, and 1,321 for unvaccinated and uninfected individuals and 11,752, and 5,050 for vaccinated and infected (Fig.1E). In comparison, we assessed the NAb and RBD-binding antibody titer in non-KTRs. Despite few cases of uninfected individuals, vaccinated and infected participants showed higher level of NAb titer than vaccinated only, infection only, or none. Interestingly, infection-only trended higher than vaccination-only group (Fig. S1E). RBD-binding antibody titer aligned the neutralization results, vaccinated and breakthrough infected individuals showed the greatest binding antibody titer (11,854 and 6,111 for D614G and JN.1, respectively) (Fig. S1F). Taken together, the results indicate that the breakthrough infection is able to boost immunity against SARS-CoV-2, without excluding the immunosuppressive individuals.

### Immunosuppressant dosage and duration of therapy are inversely correlated with antibody production

We next investigated the key determinants that impairs the antibody responses in KTRs. We firstly examined the potential effects of immunosuppressant (IS) dosage and duration of therapy on antibody production. After stratify the cohorts into non-KTR, 2 drug receivers, and 3 drug receivers, there is progressive decline of antibody titer with increased immunosuppressant dosage, with NAb titer of 158.6, 36.7, and 20 against D614G and 28.9, 34.5, and 20 against JN.1, respectively (Fig. S2A). Further, we stratify the cohorts by therapy duration, and noted treatment greater than 6 months, treatment less than 6 months, and non-KTR showed a NAb titer of 158.6, 51.8, and 20 against D614G and 28.9, 20, and 20 against JN.1, respectively (Fig. S2B). In addition, the kidney dialysis also showed similar trend, NAb titer of 158.6, 55.8, and 20 against D614G and 28.9, 33.4, and 20 against JN.1, respectively (Fig. S2C).

Moreover, we analyzed the effects of tumor-bearing and life styles on antibody production. Interestingly, the burden of tumor did not significantly jeopardize the antibody level in enrolled individuals (Fig. S2D). Lifestyles including tobacco and alcohol consumption, night owls and additional medicine taken showed similar neutralizing antibody titers to the otherwise healthy counterparts (Fig. S2E-H). Taken together, the results indicate that IS dosage and treatment duration may serve the key determinants of antibody production in organ transplanted individuals.

### Acute immunosuppressive treatment causes quantitative, rather than qualitative, impairment of immune cell function, which is reversible upon temporary treatment interruption

To mechanistically characterize how the immunosuppressant affects the overall immune system, we established a mouse model using drug combinations that mirror standard clinical practice in transplant recipients. The triple-drug regimen of a calcineurin inhibitor (Tacrolimus, T), an antimetabolite (Mycophenolate Mofetil, MMF), and a corticosteroid (Methylprednisolone, T) is the primary immunosuppressive therapy for kidney transplant patients. We therefore treated C57BL/6J mice with this triple combination (MTM), as well as double (MT) and single (M) drug regimens, to model a gradient of immunosuppressive intensity (Fig. 2A). The immunosuppressant treatment either continued till day 28 (continuous treatment, CT) or stopped on day 10 (interrupted treatment, IT). These regimens model, respectively, the steady-state immunosuppression in stable transplant patients and a hypothetical scenario where treatment is briefly paused to facilitate vaccination. On day 14, all mice received one-shot of Ad5-S.PP, and the serological immune parameters were determined on the weekly basis. Two days after IS treatment, mice were bled and 100 μL of whole blood were analyzed by flow cytometry, counts of leukocytes, T and B cells were marginally affected when compared with mock treated mice (Fig. 2B). Cell counts on day 10 in contrast showed a progressive decrease. Compared with mock group, CD45+ leukocytes, CD3+ T cell, CD4+ T cells, CD8+ T cells, and CD45R+ B cells declined by 1.32, 1.28, 1.31, 1.22, 1.38-fold in M group, by 2.81, 3.65, 3.82, 3.37, 2.89-fold in MT group, and by 5.33, 5.08, 6.07, 4.05, 8.22-fold in MTM group (Fig. 2C). The according cell frequency are less affected except those from the MTM triple treatment (Fig. S3A).

**Figure 2.**
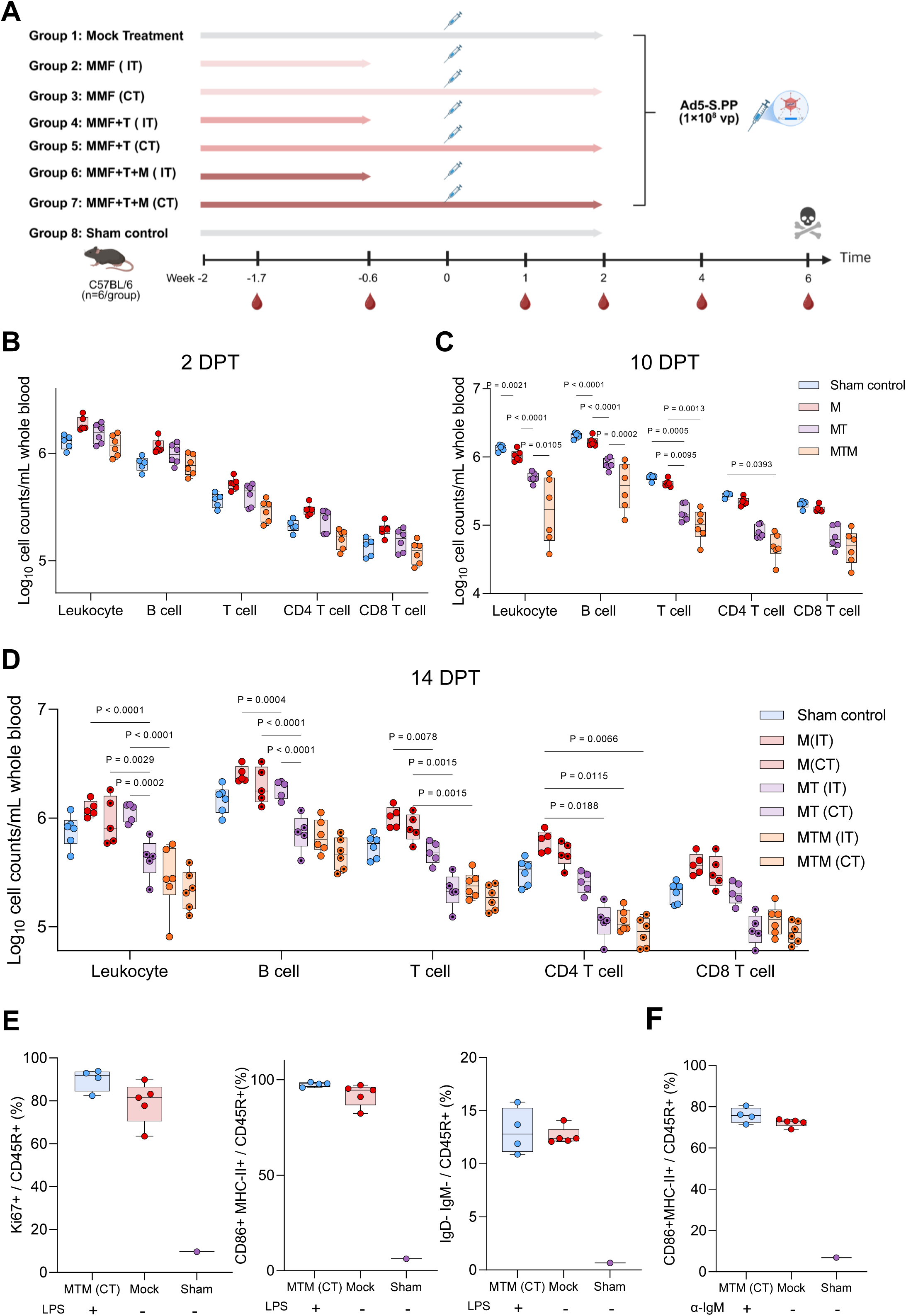
Immunosuppressants cause quantitative, reversible impairment of immune cell function. (A) Experimental timeline. C57BL/6 mice (n=6 per group) received the indicated immunosuppressant (IS) regimens via daily oral gavage: Mycophenolate Mofetil (M); MMF + Tacrolimus (MT); or MMF + T + Methylprednisolone (MTM). Treatment was either continuous (CT) for 28 days or interrupted (IT, stopped on day 10). All mice were immunized intramuscularly with Ad5-S.PP (1×10^8^ viral particles, vp) on day 14. (B-D) Absolute counts of peripheral blood immune cell subsets, as determined by flow cytometry, on day 2 (B), day 10 (C), and day 14 (D) after IS initiation. Cell populations quantified: leukocytes (CD45+), T cells (CD3+), helper T cells (CD4+), cytotoxic T cells (CD8+), and B cells (CD45R+). (E, F) Functional analysis of B cells after 10 days of triple IS treatment (MTM continuous regimen). PBMCs were stimulated for 72 hours with LPS (E) or anti-IgM antibody (F). B cell proliferation (Ki67+), activation (CD86+MHC-II+), and class-switching (IgD-IgM-) were assessed by flow cytometry. Data are presented as median ± IQR (n=6 mice/group). Statistical significance was determined by two-sided Mann-Whitney U tests.

To determine if IS treatment dynamically regulates immune cell counts, we analyzed PBMCs at day 14 post treatment (14 DPT or 4 days post-withdrawal). Although IS treatment reduced overall immune cell counts in a dose-dependent manner, the CD45+, CD3+, CD4+, CD8+, and CD45R+ cell subsets recovered almost fully after single– and double-IS withdrawal (IT groups). Interestingly, triple-IS treatment permitted only partial recovery, indicating a dose-dependent effect on immune cell recovery (Fig. 2D).

We further assessed whether the IS treatment led to functional alteration of T and B cell repertoires. PBMCs were isolated from the mice on day 10, and treated with 100 ng/ml lipopolysaccharide (LPS) or IgM antibody, the B and T cell responses were analyzed by flow cytometry. As shown in Fig. 2E-F, the proliferation (Ki67+CD45R+), activation (CD86+MHCII+CD45R+), and class switch (IgD-IgM-CD45R+) status of B cells after stimulation by LPS or IgM antibody crosslinking were comparable in mock treated and CT groups (triple drug). Similarly, IFNγ– and TNFα-specific CD8+ or CD4+ T cells after LPS stimulation were comparable in both groups (Fig. S3B-E).

Together, these data indicate that IS treatment impairs immune cell numbers without substantially affecting their function, and this numerical reduction is reversible upon treatment withdrawal.

### Acute immunosuppressive treatment causes a dose-dependent and reversible impairment of vaccine-elicited immunity

Immunosuppressant treated mice were immunized with single-shot of Ad5 S.PP at day 14, and bled at 2-, 4-, and 6-weeks post immunization for antibody assessment. Compared with vaccination only group which elicited a median RBD-specific binding IgG titer of 33,115 at week 2 post immunization, the single-, double-, and triple-IS continuous treatment led to 11.3-, 331.1-, and 331.1-fold reduced binding antibody titer (Fig. 3A, left panel). Interestingly, interrupted IS treatments recovered partially the antibody titers, particularly for double– and triple IS treatments, which recovered from lower limit of detection (<100) to 15,786, and 5,798, respectively. At weeks 4 and 6 post immunization, the antibody titers reached to an equivalent level to vaccination only group, regardless of the IS treatment regimens (Fig. 3A, middle and right panels). In align with the binding antibody titer, NAb titer against D614G and Delta demonstrated a similar trend (Fig. 3B-C). At week 2 post immunization, NAb titer was reduced by 4.97-, 4.97-, and 4.97-fold for single-, double-, and triple-IS treatment when compared with vaccination only control group, while interrupted IS treatment partially recovered the suppression, with only 2.28, 1.57, and 1.72-fold reduction for single-, double-, and triple-IS treatment. At week 4 and 6 post immunization, the continuous IS treatment groups generally showed antibody titers not significantly different to the interrupted groups, implicating a gradual recovered antibody production after IS treatment retraction. NAb against Delta variant similarly suppressed at week 2 post immunization, and gradually recovered from week 4 to week 6 after IS treatment retraction, despite a generally lower NAb titer were elicited than ancestral D614 (Fig. 3C). To identify immune parameters linked to the antibody response in immunocompromised individuals, we correlated immune cell counts with anti-spike IgG titers. We found that both B and T cell counts positively correlated with spike-specific IgG levels (Fig. 3D). The positive correlation between B and T cell counts themselves suggests that the immunosuppressive therapy broadly depleted these key lymphocyte populations.

**Figure 3.**
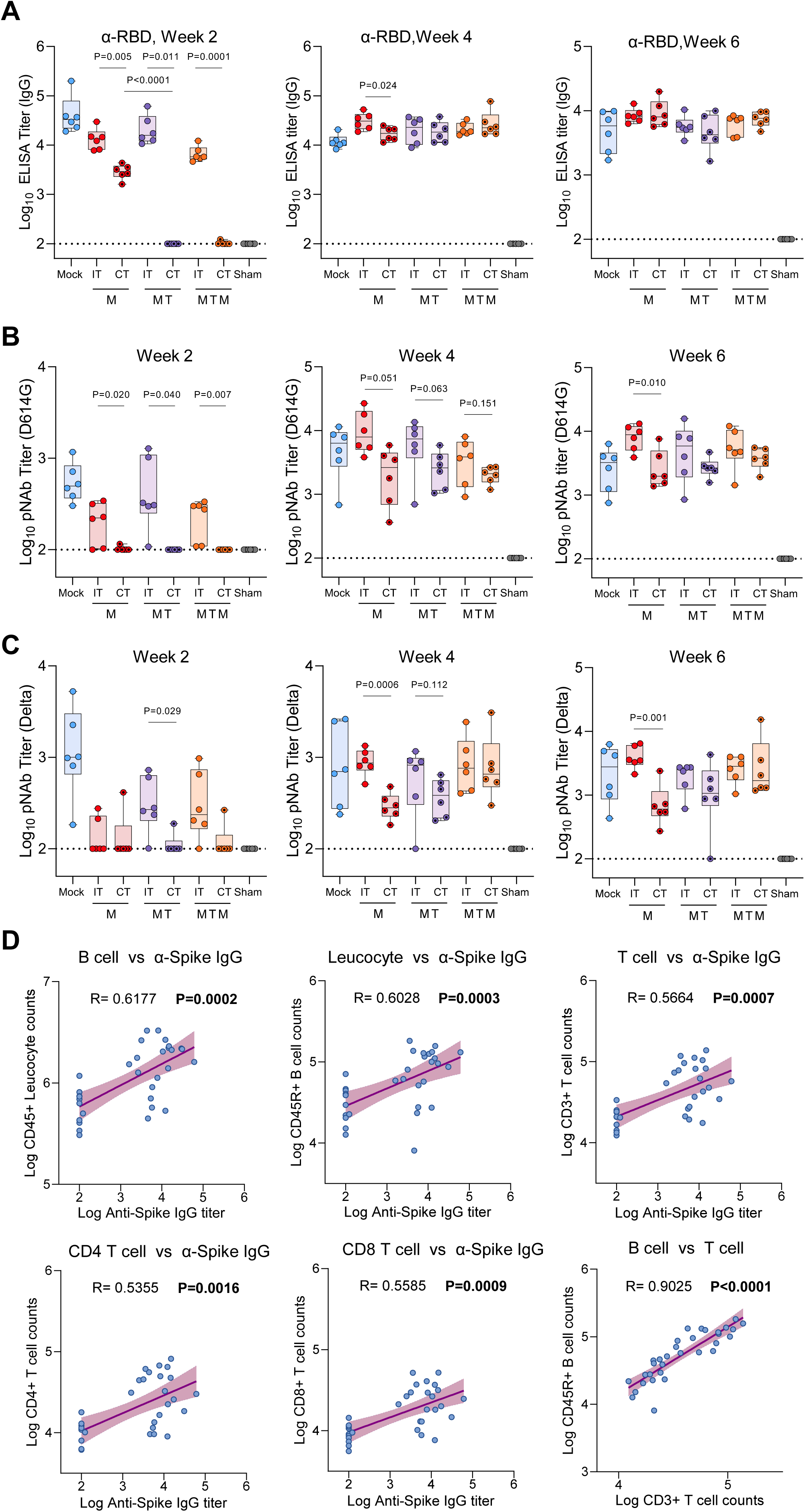
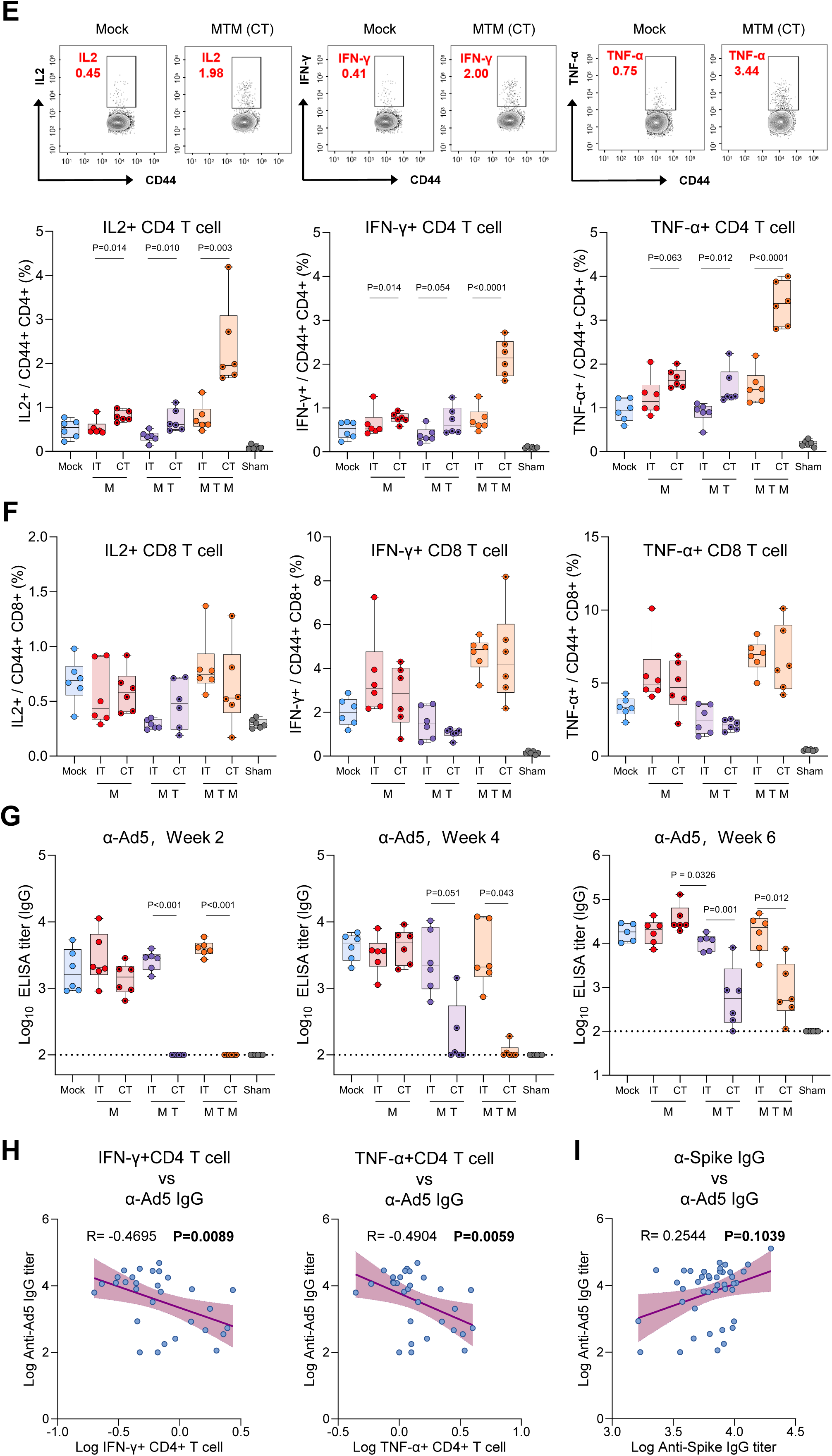
Immunosuppressants cause a dose-dependent and reversible impairment of vaccine-elicited immunity. (A) Serum RBD-specific IgG titers against the wild-type (WT) RBD at weeks 2, 4, and 6 post-immunization with Ad5-S.PP. Mice were treated with the indicated IS regimens as described in Fig. 3A. (B, C) Neutralizing antibody (NAb) titers against the D614G (B) and Delta (C) variants at the indicated time points. (D) Correlation analysis between immune cell counts (measured at day 14 post-IS initiation) and spike-specific IgG titers (measured at week 2 post-vaccination). Spearman correlation coefficient (R) and P value are shown for each analysis. (E, F) Intracellular cytokine staining (ICS) analysis of splenic T cells at week 6 post-immunization. Representative flow cytometry plots (top) and quantification (bottom) of cytokine-producing CD4+ (E) and CD8+ (F) T cells after stimulation with SARS-CoV-2 spike peptide pools. (G) Serum anti-Ad5 vector IgG titers at weeks 2, 4, and 6 post-immunization, as determined by ELISA. (H) Negative correlation between the frequency of IFN-γ+ or TNF-α+ CD4+ T cells (from E) and the anti-Ad5 IgG titer at week 6. (I) Correlation between anti-spike IgG and anti-Ad5 IgG titers at week 6. For (A-C, E-G), data are presented as median ± IQR (n=6 mice/group). Statistical significance was determined by two-sided Mann-Whitney U tests for comparisons between groups and by Spearman correlation test for (D, H, I). Dashed lines indicate the lower limit of quantification (LLOQ: 100 for ELISA, 20 for NAb). IS, immunosuppressant; CT, continuous treatment; IT, interrupted treatment.

The gradual recovery of immune suppression following IS withdrawal was further supported by the analysis of terminal B and T cell responses. ELISPOT assays for IgG-specific B cells and IL-4-specific T cells showed no significant differences among the groups (Fig. S4A-B). Similarly, ICS assays revealed no overall difference in CD8+ or CD4+ T cell responses across the various IS treatment regimens (Fig. 3E-F). Notably, however, mice subjected to continuous IS treatment, particularly the triple-drug regimen, exhibited significantly higher CD4+ T cell responses and a trend toward higher CD8+ T cell responses. We hypothesized that anti-Ad5 immunity may have been suppressed in the continuous treatment (CT) groups, thereby creating a more favorable environment for T cell responses under IS. To test this, we measured anti-Ad5 IgG in mouse serum and observed strong inhibition in the two– and three-drug CT groups from week 2 to week 6 (Fig. 3G).

Correlation analysis demonstrated a strong inverse relationship between spike-specific CD4+ T cell responses (IFNγ+ and TNFα+ T cell) and anti-Ad5 IgG titers (Fig. 3H). This suggests that anti-Ad5 antibody likely exerts an inhibitory effect on T cell responses, and that IS treatment may unexpectedly enhance T cell activity by suppressing this antibody response. Interestingly, anti-Ad5 IgG titers were trending positively correlated with anti-Spike IgG titers (Fig. 3I). The stark contrast in how humoral and cellular immune responses relate to anti-Ad5 titers implies that IS treatment modulates these two arms of immunity through intrinsically distinct mechanisms.

Collectively, our results suggest that the immunosuppressive effect of the three tested drugs is mediated primarily by a dose-dependent reduction in immune cell numbers, not by specific pathway inhibition. Essentially, treatment discontinuation subsequently permitted the recovery of antibody titers.

### Adenoviral vector-based vaccines elicited enhanced immunogenicity in immunosuppressed murine models

To evaluate the impact of vaccine modality on immunogenicity under immunosuppressive therapy, we performed a comparative analysis of the Ad5-vectored vaccine (Ad5-S.PP) and a protein-based vaccine (S.PP). The experimental design (Fig. 4A) featured identical continuous (CT) and interrupted (IT) IS regimens, with S.PP immunization administered at weeks 0 and 1.4. Continuous IS treatment suppressed anti-RBD (D614G) antibody titers in both vaccine groups at two weeks post-immunization. In contrast, withdrawal of IS elicited a 57.9-fold increase in antibody titers specifically in the Ad5 S.PP group (Fig. 4B). At week 4 post immunization, the binding antibody titer showed constant increase, while the titer remained low in IS continuous group, instead IgG titer in IT group reached a level equivalent to the untreated group. At week 6 post immunization, the protein vaccinated groups kept increasing, a level even higher than Ad5.S.PP groups, however, the continuous treated group still showed a level lower than interrupted group. The NAb titer against both D614G and Delta showed the similar trend to the IgG titer, showing a partial recovery in IT groups (Fig.4C). Again, the NAb titer were fully recovered for Ad5 S.PP starting at week 4 post immunization and maintained at least to week 6 post immunization, while S.PP immunized mice showed a level still significantly reduced in IS treated group even at week 6 post immunization. Notably, the NAb against Delta variant elicited by S.PP under continuous triple-IS (CT) treatment remained below the limit of detection throughout the experiment. In stark contrast, Ad5-S.PP elicited high and sustained NAb titers (Fig. 4D, right panel), underscoring the platform’s superiority in inducing functional antibodies under potent immunosuppression. Given that both S.PP and Ad5-S.PP robustly induced IgG, yet only Ad5-S.PP generated robust NAbs against variant, these results strongly implicate that the Ad5 platform is superior in driving antibody affinity maturation even under potent immunosuppressive therapy.

**Figure 4.**
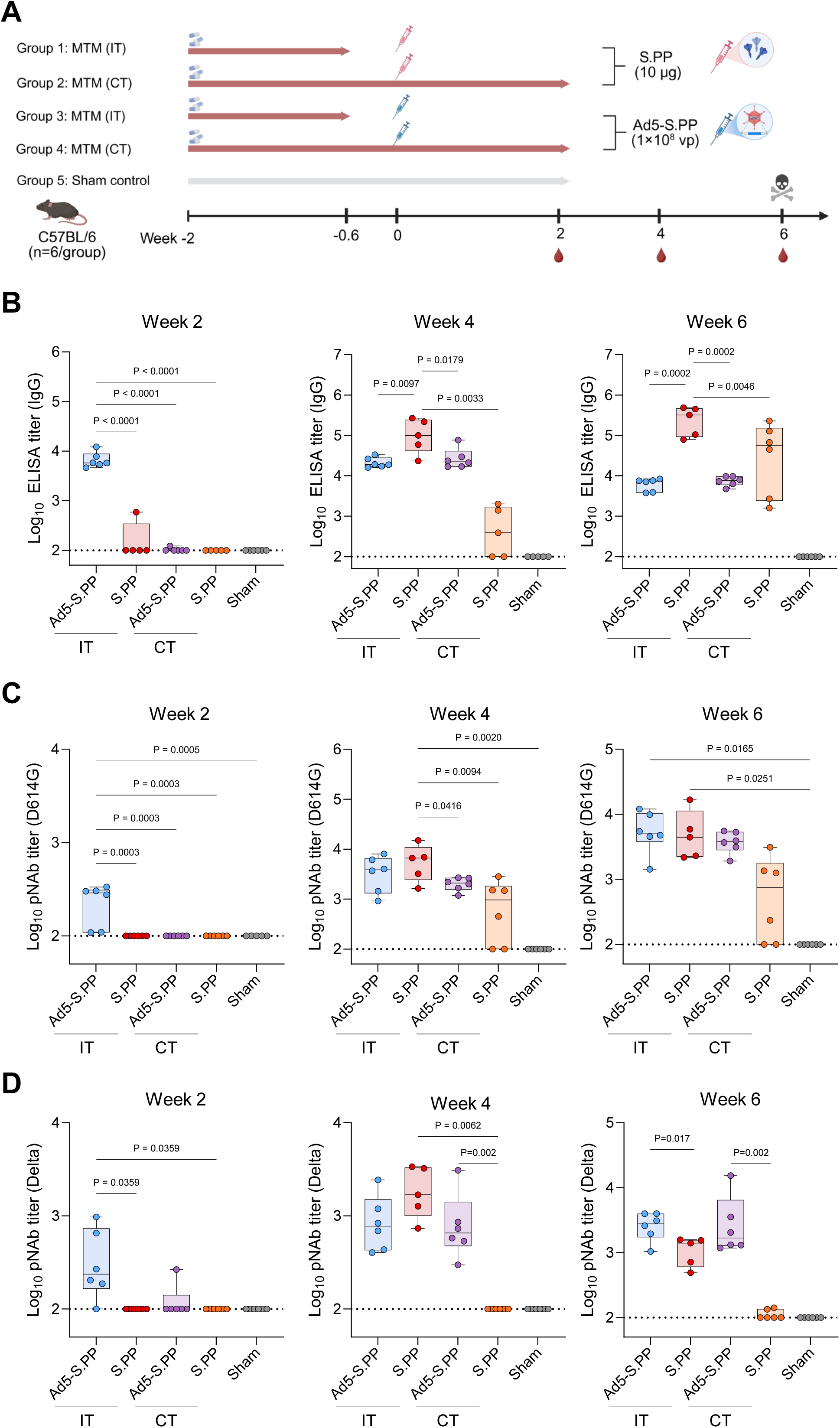
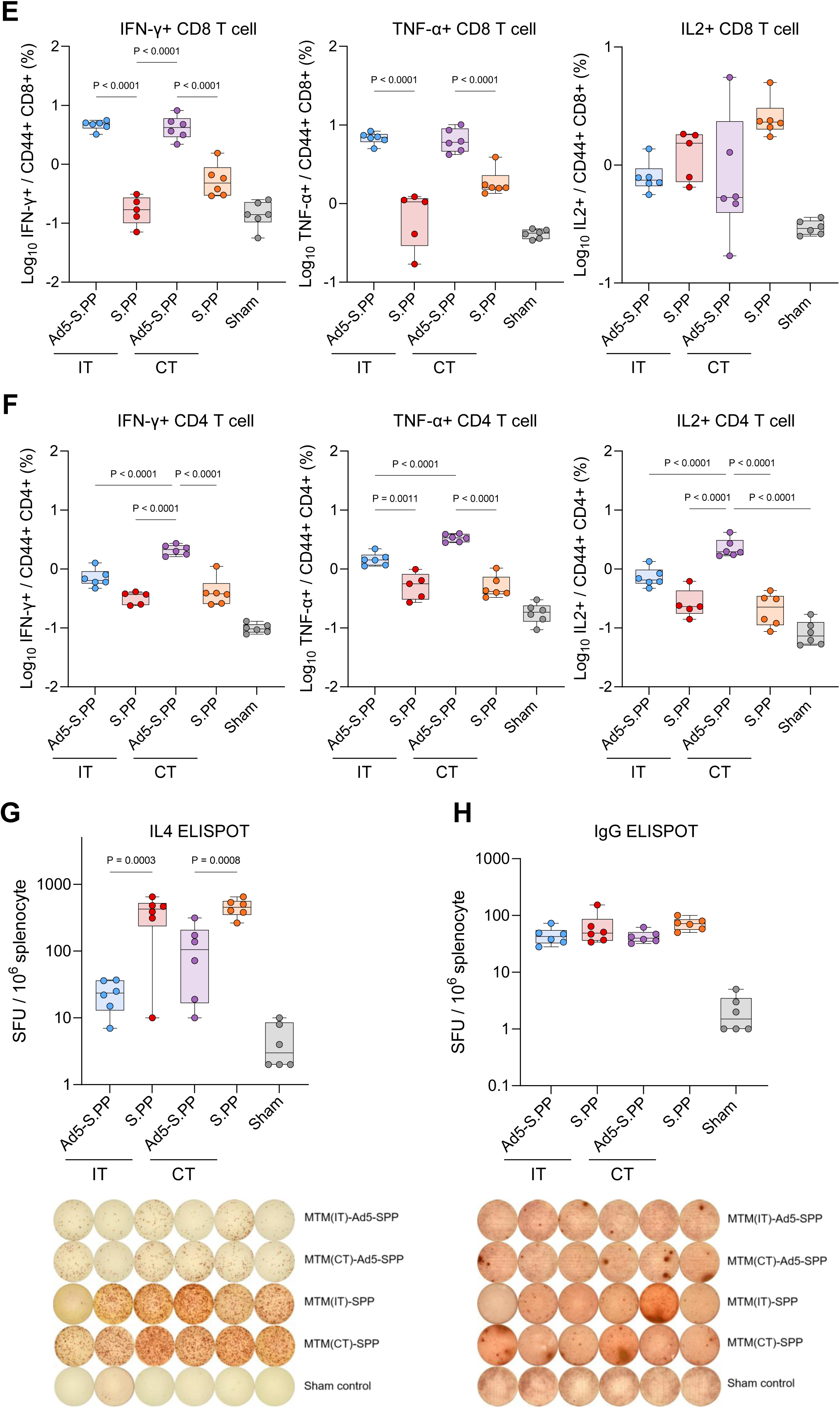
Adenoviral vector vaccines elicit enhanced immunogenicity in immunosuppressed mice. (A) Experimental timeline. Mice were administered the triple-drug IS regimen (MTM) and immunized with either Ad5-S.PP (1×10^8^ vp, single dose on day 0) or protein S.PP (10 µg adjuvanted with Alum, two doses on days 0 and 28). The interrupted therapy (IT) group stopped IS treatment on day 10. (B) Serum RBD-specific IgG titers against WT-RBD at weeks 2, 4, and 6 after the initial immunization. (C, D) Neutralizing antibody (NAb) titers against the D614G (C) and Delta (D) variants at the indicated time points. (E, F) Quantification of cytokine-producing CD4+ (E) and CD8+ (F) T cells from splenocytes at week 6, assessed by ICS after spike peptide pool stimulation. (G) Frequency of IL-4-secreting T cells measured by ELISpot at week 6. (H) Number of spike-specific IgG-secreting B cells measured by ELISpot at week 6. For (B-H), data are presented as median ± IQR (n=5-6 mice/group). Statistical significance was determined by two-sided Mann-Whitney U tests for comparisons between the two vaccine platforms under the same treatment condition (NT, IT, or CT). Dashed lines indicate the LLOQ. NT, no treatment; IT, interrupted treatment; CT, continuous treatment.

Moreover, the Ad5-S.PP vaccine induces a significantly stronger CD8+ and Th1-biased CD4+ T cell response (Fig. 4E-F), along with a lower frequency of Th2-biased IL-4+ T cells (Fig. 4G). Interestingly, IgG+ B cell ELISpot analysis revealed comparable numbers of antibody-secreting B cells in S.PP treated mice (Fig. 4H), implicating that the differences between adenovirus– and protein-based immunization lies in antibody production rather than in the quantity of B cells generated.

### Adenoviral vector-based vaccines sustained high and durable antigen expression even during concurrent immunosuppressive therapy

To investigate the mechanism underlying the enhanced immunogenicity of the Ad5-based vaccine under IS treatment, we employed a secreted embryonic alkaline phosphatase (SEAP) reporter system ^33^. This method provided a sensitive, enzymatic readout of transgene expression in serum, circumventing the technical challenges of directly measuring Spike protein over time. As outlined in Fig. 5A, C57BL/6 mice were pre-treated with IS for eight days before immunization with Ad5-SEAP (day 0). SEAP activity and IgG titers against both the Ad5 vector and the SEAP protein were monitored longitudinally. Intriguingly, SEAP activity in the mock-treated group peaked at day 2 and gradually declined to baseline levels by day 10. In contrast, the CT group exhibited similarly high SEAP levels at day 2, but this activity increased further until day 10, only beginning to decline after day 14 and remaining elevated through day 21. The IT group also showed high initial SEAP secretion until day 7, after which it underwent a sharp decline to levels comparable to the mock-treated group (Fig. 5B). Analysis of the area under the curve (AUC) confirmed that Ad5-SEAP expression was highest under CT conditions over the entire 21-day period (Fig. 5C).

**Figure 5.**
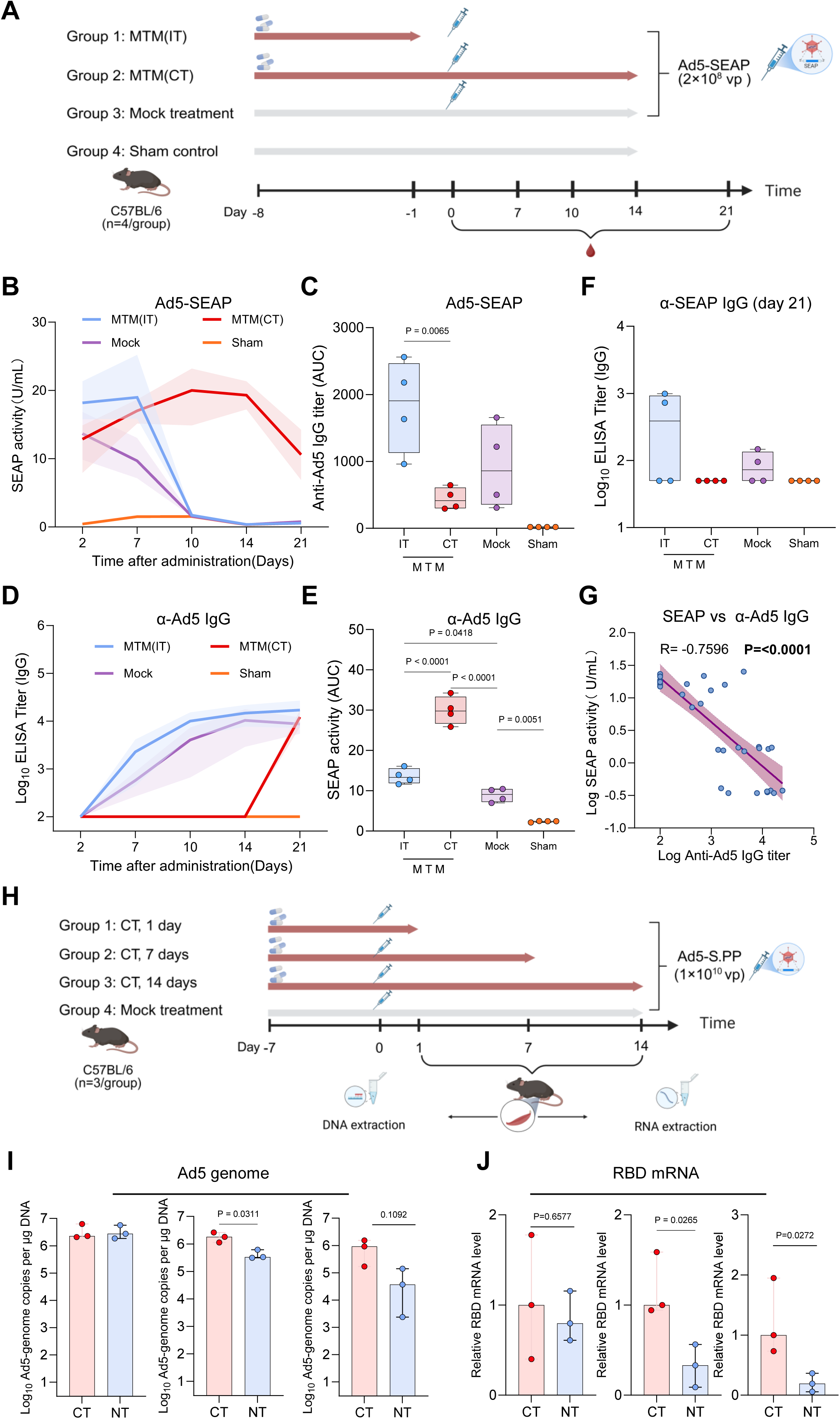
Adenoviral vector vaccines sustain antigen expression under immunosuppressive therapy. (A) Experimental schema. Mice were pre-treated with the triple-drug IS regimen (MTM) for 8 days before intramuscular immunization with Ad5-SEAP (1×10^9^ vp, day 0). (B) Longitudinal monitoring of serum SEAP activity, a surrogate for transgene expression levels. (C) Area under the curve (AUC) analysis of the SEAP activity data shown in (B). (D) Longitudinal measurement of serum anti-Ad5 IgG titers. (E) AUC analysis of the anti-Ad5 IgG data shown in (D). (F) Serum anti-SEAP IgG titers at the endpoint (day 21). (G) Negative correlation between serum SEAP activity and anti-Ad5 IgG titer at day 21. Spearman correlation coefficient (R) and P value are shown. (H) Schema for tissue collection to analyze vector persistence and transgene expression. (I) Quantification of Ad5 hexon DNA copies in the quadriceps muscle at the indicated days post-Ad5-S.PP immunization, with or without continuous IS (CT) treatment. Data are normalized to the mock-treated group. (J) Quantification of spike RBD cDNA transcripts in the quadriceps muscle, normalized to the mock-treated group. For (B, D), data are presented as mean ± SEM (n=5-6 mice/group). For (C, E, F, I, J), data are presented as median ± IQR. Statistical significance was determined by two-sided Mann-Whitney U tests (C, E, F, I, J) or Spearman correlation test (G).

We hypothesized that the decline in the IT group was linked to the induction of antibodies against the Ad5 vector or the SEAP protein. Indeed, anti-Ad5 IgG titers in the mock-treated group began rising from day 2 onward, whereas continuous IS treatment markedly slowed this anti-Ad5 antibody response (Fig. 5D-E). Anti-SEAP antibodies were detectable but low overall, with a similar trend toward higher titers in the IT group (Fig. 5F). A strong negative correlation was observed between anti-Ad5 antibody titers and SEAP activity (Fig. 5G), suggesting that IS treatment may enhance and sustain transgene expression by suppressing the neutralizing anti-vector antibody response. This elevated expression in the CT condition was not antigen-specific, as the same phenotype was observed with Ad5-Luc vectors (Fig. S5D-E).

This durable expression has been previously attributed to the long-term episomal persistence of adenoviral vectors ^34^. To test this, we longitudinally quantified Ad5 genome copies and RBD transcripts in mouse quadriceps with or without continuous IS treatment (Fig. 5H). Consistent with the expression data, CT led to significantly higher Ad5 hexon DNA copies in muscle tissue at days 7 and 14 post-immunization (Fig 5I, S5A and S5C). In alignment with the Ad5 genomic DNA data, spike RBD cDNA levels were also higher in the CT groups (Fig. 5J and S5B).

Together, these data demonstrate that by blunting pre-existing immunity to the Ad5 vector, IS treatment permits prolonged viral persistence and sustained transgene expression, ultimately creating a favorable profile for vaccine immunogenicity under specific conditions.

### Ad5 vectors reprogram lipid metabolism and elicit stronger immune responses than protein-based modality

To determine whether the Ad5 vector mobilizes immune programs distinct from those of an adjuvanted protein vaccine, we performed bulk RNA-seq on draining lymph nodes from immunized mice (Fig. 6A). The Ad5-S.PP platform induced a greater number of differentially expressed genes (DEGs) under continuous immunosuppression (CT) compared to the protein vaccine, while their responses were more similar in the absence of IS (NT) or after its withdrawal (IT) (Fig. 6B).

**Figure 6.**
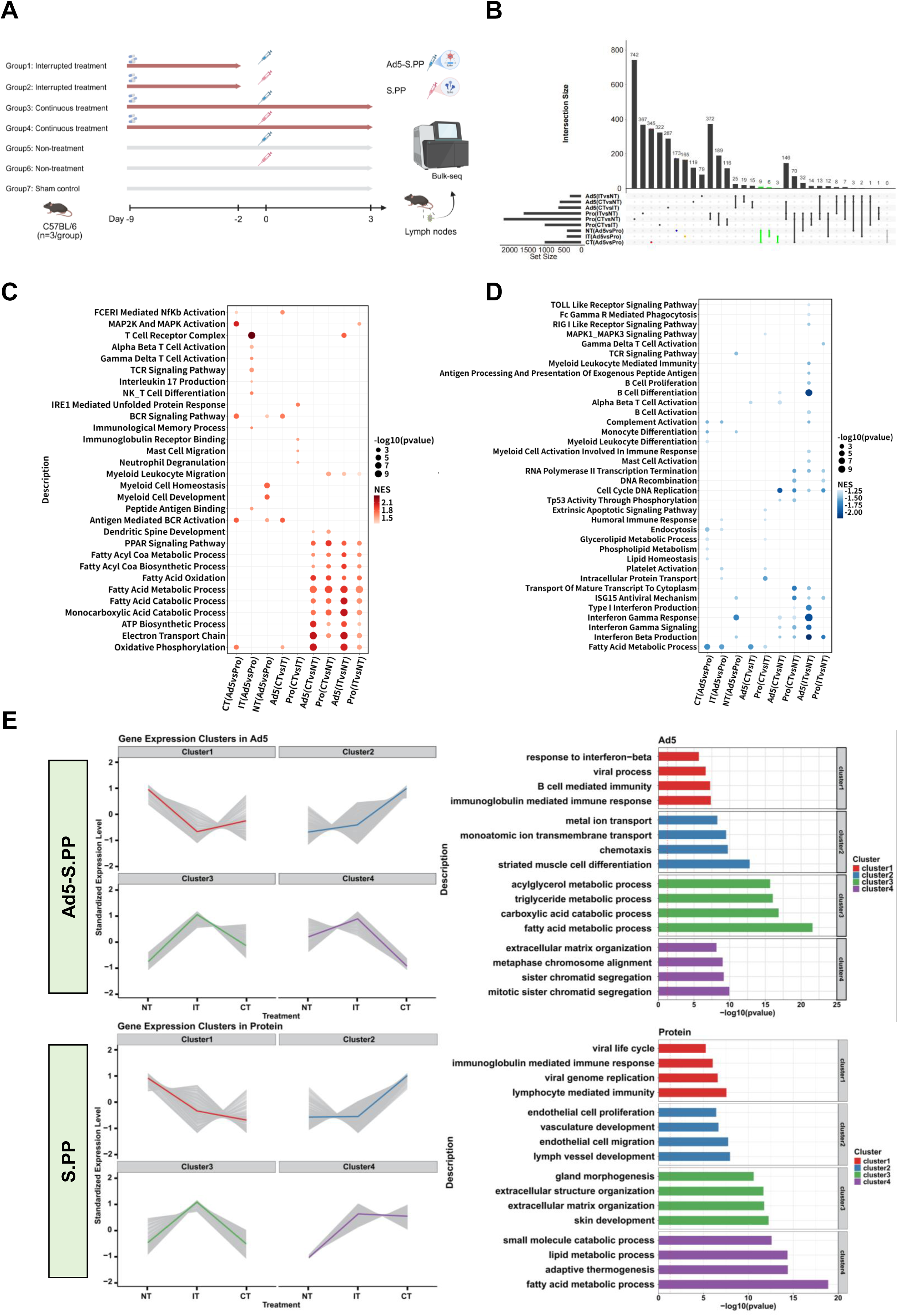
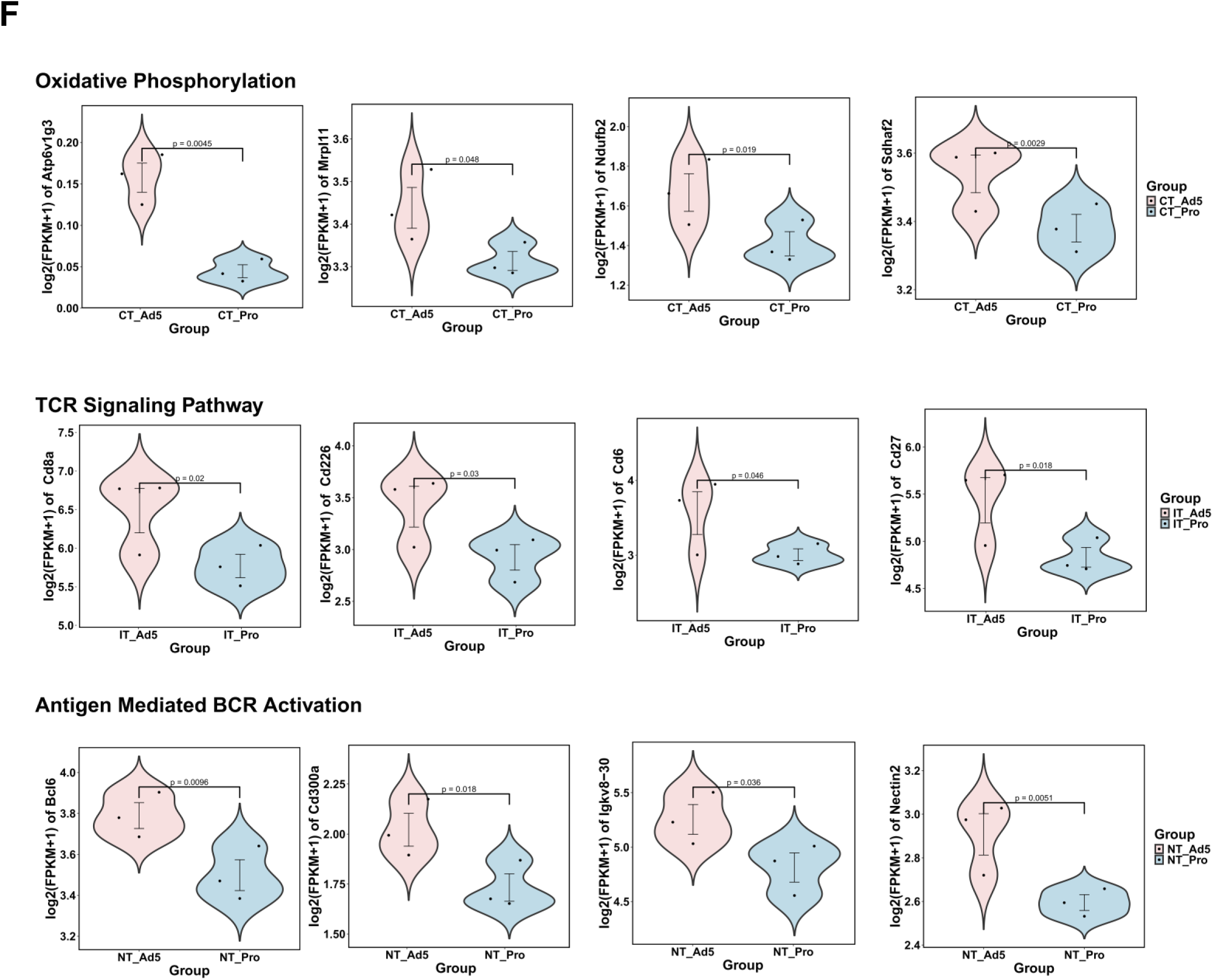
Lymph node transcriptomics reveals distinct vaccine-specific pathways. (A) Experimental design. Draining lymph nodes were harvested from mice (n=3 per group) 3 days after immunization with Ad5-S.PP or S.PP under the indicated IS conditions (NT, IT, CT) for RNA-sequencing analysis. (B) UpSet plot illustrating the number of unique and shared differentially expressed genes (DEGs) across the specified comparisons. Ad5, Ad5-S.PP; Pro, S.PP. (C, D) Gene Set Enrichment Analysis (GSEA) for comparisons between vaccine platforms under different IS conditions. Circle size represents the –log10 (adjusted P value), and color indicates the normalized enrichment score (NES). (C) Pathways positively enriched in the Ad5-S.PP group. (D) Pathways positively enriched in the S.PP group. (E) c-Means clustering of 4,829 significant DEGs across all groups, revealing four distinct co-expression clusters (C1-C4). The line plots show the expression pattern (z-score) of genes within each cluster across the different experimental groups. Selected significantly enriched biological pathways for each vaccine-specific cluster are listed on the right. (F) Violin plots showing the normalized expression of selected oxidative phosphorylation (upper panel), T cell immune response (middle panel), antigen mediated BCR activation (lower panel) genes from the Ad5-S.PP and S.PP groups under different IS treatments. For (E, F), gene expression is presented as z-score normalized counts.

Pathway enrichment analysis of these DEGs uncovered fundamentally different mechanisms of immune activation. In the absence of IS, Ad5-S.PP strongly enriched signatures for B cell receptor (BCR) signaling and myeloid cell development (Fig. 6C). Critically, under continuous IS, the most clinically relevant condition for transplant patients, Ad5-S.PP uniquely and significantly enriched pathways for oxidative phosphorylation and lipid metabolism (Fig. 6C). This metabolic reprogramming was absent in the protein vaccine group, which instead activated pathways related to endocytosis and general lipid metabolism, likely a reflection of its alum adjuvant (Fig. 6D).

We further dissected these platform-specific responses using c-Means clustering, which grouped 4,829 significant DEGs into four coherent expression patterns (Fig. 6E). This analysis confirmed that immune-related genes (Cluster 1) were suppressed under CT in both groups. However, the responses to each vaccine diverged markedly. For Ad5-S.PP, the CT condition specifically upregulated a gene cluster (Cluster 2) involved in ion transport and chemotaxis, while the IT condition (treatment withdrawal) robustly activated a distinct cluster (Cluster 3) governing acylglycerol and triglyceride metabolism (Fig. 6E, upper panel). This metabolic signature was not observed with the protein vaccine, underscoring a unique ability of the adenovirus platform to rewire immunometabolism.

Examination of individual genes solidified these findings. Ad5-S.PP immunization under CT led to the pronounced upregulation of key oxidative phosphorylation genes (e.g., *Atp6v1g3, Ndufb2*), and the IT condition triggered a sharp increase in T-cell receptor-related genes (e.g., *Cd8a, Cd27*) (Fig. 6F, S6A). Consistently, immune cell deconvolution analysis estimated a higher abundance of T cells in the Ad5-S.PP IT group (Fig. S6B).

Collectively, these transcriptomic data demonstrate that the Ad5 vector, unlike the protein vaccine, activates a robust immunometabolic program under immunosuppression. The enrichment of oxidative phosphorylation and lipid metabolic pathways likely provides the essential bioenergetic and biosynthetic substrates required for the enhanced T and B cell activation and antibody affinity maturation we observed, thereby conferring a critical advantage in this challenging immune environment.

## DISCUSSION

This study demonstrates that immunosuppression, particularly in kidney transplant recipients, severely impairs humoral immunity following COVID-19 vaccination, and establishes that this defect is both dose-dependent and functionally reversible. We further establish that adenoviral vector vaccines are able to sustain immunogenicity under IS treatment, an advantage we mechanistically link to sustained antigen presentation and a unique capacity for immune metabolic reprogramming. These findings underscore that the choice of vaccine platform is a critical, modifiable factor for protecting this vulnerable population.

Our clinical observations align with and extend a substantial body of literature documenting poor vaccine responses in transplant recipients. Numerous studies, including from our group, have established that solid organ transplant recipients on multi-drug regimens mount inadequate serological responses to mRNA and inactivated COVID-19 vaccines (7-14). Here we identified the dosage and duration of immunosuppressive therapy as the most critical parameters (Fig. S2A-B). The first-line regimen of organ transplants, comprising an antimetabolite, a calcineurin inhibitor, and a corticosteroid, acts by globally targeting proliferating lymphocytes ^35^. Consistent with some clinical observations ^36–38^, our controlled murine models provide direct experimental proof that this acute immunosuppression-induced deficit is reversible. The rapid recovery of antibody production upon treatment interruption, coupled with the comparable antibody maturation profiles in KTRs and non-KTRs, indicates a primarily quantitative but not qualitative impairment. This suggests that short courses of immunosuppressive regimens cause dose-dependent depletion of immune cells, particularly B cells (Fig. 2C-D, 3D), without inflicting permanent functional damage (Fig. 2E-F, S3, S4). Consequently, upon the alleviation of this suppressive pressure, the reconstituted lymphocyte pool can mount qualitatively normal responses. This highlights the tantalizing potential of strategically timed treatment breaks to enhance vaccine immunogenicity, provided the risks to graft rejection are carefully managed.

The relative superior performance of the adenoviral vector platform under immunosuppression represents a significant advance. We mechanistically linked this advantage to a multifaceted mode of action. First, we discovered that immunosuppression paradoxically enhances the durability of vaccine itself. By blunting the anti-vector antibody response (Fig. 3G, 5D-E), continuous IS treatment permitted prolonged viral persistence and sustained antigen expression (Fig. 5B-C, I-J). This extended antigen availability appears to compensate for the immunosuppressive environment by providing a persistent antigenic stimulus, which is crucial for initiating and maintaining immune responses in a lymphopenic host.

Second, and perhaps more profoundly, transcriptomic analysis revealed that the adenoviral vector, unlike the protein vaccine, engages distinct and more resilient immune pathways. Under continuous immunosuppression, Ad5-S.PP uniquely enriched pathways related to oxidative phosphorylation and lipid metabolism (Fig. 6C, E, F). This coordinated immunometabolic reprogramming is intrinsically linked to immune cell activation, proliferation, and antibody affinity maturation, as it provides the necessary bioenergetic and biosynthetic substrates. This intrinsic adjuvantity may allow adenoviral vectors to “break through” the immunosuppressive barrier more effectively than protein antigens, which rely more heavily on exogenous alum adjuvant that may be susceptible to suppression. The functional outcome of this metabolic advantage is clear: despite eliciting comparable total IgG, Ad5-vectored vaccines induced significantly higher levels of cross-reactive neutralizing antibodies than the protein-based platform (Fig. 4B-D), indicating not only greater quantity but also superior quality of the humoral response.

Despite these insights, our study has limitations. The human cohort, while informative, was observational, and findings related to treatment interruption require validation in a controlled clinical trial setting to assess both efficacy and the associated risk of graft rejection. Our murine model, while allowing for precise mechanistic dissection, utilizes a specific short course of triple-drug regimen and may not fully recapitulate the complex and heterogeneous immune status of long-term transplant recipients. Furthermore, the human cohort included only a small number of individuals receiving adenoviral vector vaccines (n=3), limiting the statistical power for direct platform comparison in the clinical setting. The analysis of cellular immunity in the human cohort was also limited, and the long-term durability of the immune responses elicited by different platforms remains to be determined. Finally, the potential impact of pre-existing adenovirus immunity, a known concern for vector-based vaccines, was minimized in our model but warrants careful consideration in broader human populations.

Despite these limitations, the significance of our findings is substantial. Immunocompromised individuals remain at high risk for severe outcomes from infectious diseases and are often left behind by conventional vaccine approaches. Our data provide a scientific rationale for moving beyond a one-size-fits-all vaccination strategy. They underscore that vaccine modality is a critical variable, with adenoviral vectors emerging as a highly promising platform due to their robust, sustained, and metabolically enhanced immunogenicity, even under significant immunosuppressive pressure. Future efforts should focus on engineering next-generation viral vectors that further minimize pre-existing immunity interference while maximizing these beneficial innate immune and metabolic activation pathways. By rationally tailoring vaccine platforms to the unique immunological landscape of the immunosuppressed host, we can take a crucial step toward providing equitable and effective protection for this vulnerable population.

## MATERIALS AND METHODS

### Clinical study design and participant characteristics

This study included kidney transplant recipients (KTRs) and a control cohort of non-transplant recipients (Non-KTRs) recruited from the Air Force Military Medical University. All participants were at least 18 years of age and provided written informed consent. The study was approved by the Ethics Committee of the First Affiliated Hospital of the Air Force Medical University (Registration No. KY20232379-C-1). All patients had completed a three-dose SARS-CoV-2 vaccination series between May 2021 and December 2022. The majority of participants (n =129, 97.7%) received an inactivated vaccine, while a minority (n=3, 2.3%) received an Ad5-nCoV vector vaccine. Demographic and clinical characteristics of the KTR and Non-KTR populations, including subgroups based on immunosuppressant regimen and dialysis duration, are summarized in Table 1. Peripheral blood was collected from KTR and Non-KTR recipients in December 2023 and August 2024 for the isolation of peripheral blood mononuclear cells (PBMCs) and serum.

### Blood processing and serum isolation

Peripheral venous blood (5–10 mL) was collected into serum separation tubes under aseptic conditions and allowed to clot at room temperature for 30–60 minutes. Samples were centrifuged at 1,500–2,000 × g for 10 minutes. The resulting serum was carefully transferred into sterile tubes, aliquoted into 1.0 mL volumes to minimize freeze-thaw cycles, and heat-inactivated at 56°C for 30 minutes. All serum aliquots were stored at –80°C until analysis.

### Enzyme-Linked Immunosorbent Assay (ELISA)

SARS-CoV-2-specific antibodies were quantified by ELISA. Briefly, 96-well plates were coated with 1.5 μg/mL of SARS-CoV-2 receptor-binding domain (RBD) or nucleocapsid (N) protein in PBS and incubated overnight at 4°C. Plates were washed once with wash buffer (0.05% Tween-20 in PBS) and blocked with 200 μL of blocking buffer (5% skim milk in PBS) per well for 2 hours at room temperature. After discarding the block solution, serially diluted serum samples (3-fold dilutions) were added to the wells and incubated for 2 hours at room temperature.

Following three washes, plates were incubated with horseradish peroxidase (HRP)-conjugated goat anti-human IgG (0.25 μg/mL; Sino Biological) for 1 hour at room temperature. Plates were washed again, and 100 µL/well of TMB substrate (Solarbio) was added. The reaction was stopped with 50 µL/well of 1 N H SO, and absorbance was measured at 450 nm using a microplate reader (BioTek Synergy H1). Endpoint titers were defined as the highest serum dilution that yielded an optical density (OD) greater than two times the background signal. The lower limit of quantification (LLOQ) for the ELISA was a titer of 100.

### Pseudovirus neutralization assay

SARS-CoV-2 spike pseudotyped lentiviruses were generated by co-transfecting HEK293T cells with three plasmids: (i) a plasmid encoding a codon-optimized, C-terminally truncated SARS-CoV-2 spike protein, (ii) the psPAX2 packaging plasmid (Addgene), and (iii) a lentiviral transfer vector expressing a luciferase reporter. Transfections were performed using polyethylenimine (PEI) in Opti-MEM. Pseudovirus-containing supernatants were harvested 48 hours post-transfection, filtered through a 0.45 μm membrane, and stored at −80°C.

For neutralization assays, HEK293T cells stably expressing human ACE2 (hACE2) were seeded in 96-well plates at 2×10 cells per well. Serum samples were serially diluted (3-fold) and mixed with an equal volume of pseudovirus. The mixture was incubated at 37°C for 1 hour before being added to the HEK293T-hACE2 cells. Plates were centrifuged at 1,680 × g for 20 minutes and then incubated at 37°C with 5% CO for 36–48 hours. Luciferase activity was measured using a Firefly Luciferase Reporter Gene Assay Kit (Beyotime) and quantified on a microplate reader (BioTek Synergy H1). The 50% neutralization titer (NT50) was defined as the serum dilution that resulted in a 50% reduction in relative light units compared to virus control wells. The LLOQ for the pseudovirus neutralization assay was an NT50 of 20.

### Antigenic cartography

Antigenic cartography was performed using pseudovirus neutralization titers from recipient sera. Maps were constructed with the Cartography server (acmacs-web.antigenic-cartography.org). SARS-CoV-2 antigens are represented as circles and sera as squares. Each grid square corresponds to one antigenic unit, equivalent to a 2-fold change in neutralization titer. The antigenic distances among the Prototype, Omicron BF.7, XBB, EG.5.1, and JN.1 variants were calculated and converted to fold-differences.

### Mouse immunosuppression

Six– to eight-week-old female C57BL/6J mice were obtained from Vital River Laboratory Animal Technology Co., Ltd. and maintained under specific pathogen-free conditions at the Guangzhou Laboratory. All animal experiments were approved by the Institutional Animal Care and Use Committee (IACUC) of Guangzhou Laboratory. To model pharmacological immunosuppression, mice were randomly assigned to four groups receiving daily oral gavage of the following regimens:

Vehicle Control; MMF: Mycophenolate mofetil (100 mg/kg/day); MMF+T: MMF (100 mg/kg/day) + Tacrolimus (6 mg/kg/day); MMF+T+M: MMF (100 mg/kg/day) + Tacrolimus (6 mg/kg/day) + Methylprednisolone (20 mg/kg/day). Immunosuppressants were dissolved in a vehicle of 10% DMSO, 40% PEG300, 5% Tween-80, and 45% saline. Each immunosuppression group was further divided into two treatment duration cohorts: continuous treatment for 28 days or interrupted treatment for 10 days.

### Mouse immunization

Mice were immunized via intramuscular injection under brief isoflurane anesthesia. They received either 1×10 viral particles (vp) of Ad5-Spike (CanSino Biologics Inc.) on day 0, or 10 μg of SARS-CoV-2 spike protein (with alum adjuvant) on days 0 and 28. Peripheral blood was collected at multiple time points (e.g., days 2, 10, 14, 28, 42, and 56) for serological analysis. Mice were sacrificed at designated endpoints for the collection of spleens and other tissues.

### Flow cytometric analysis of immune cells

PBMC immunophenotyping: Mouse peripheral blood was collected into EDTA-coated tubes, and PBMCs were isolated using ACK lysis buffer. Cells were stained with fluorochrome-conjugated antibodies against CD3, CD4, CD8, CD45, and CD45R for 30 minutes at 4°C. Data were acquired on a Sony ID7000 flow cytometer and analyzed with FlowJo software.

Intracellular Cytokine Staining (ICS): Splenocytes were stimulated for 1 hour with overlapping peptide pools spanning the SARS-CoV-2 WA1/2020 spike protein (2 μg/mL), followed by a 8-hour incubation in the presence of GolgiStop and GolgiPlug. Cells were surface-stained for CD3, CD4, CD8, CD44, and CD62L, then fixed, permeabilized, and stained intracellularly for IL-2, TNF, and IFN-γ. Data were acquired on an Agilent Novocyte Advanteon flow cytometer.

B cell activation and proliferation: PBMCs were stimulated for 72 hours with LPS (1 μg/mL), IL-4 (50 ng/mL), or anti-IgM antibody (5 μg/mL). Cells were then stained for viability and surface markers (CD45R, CD86, MHC-II, IgD, IgM), followed by intracellular staining for Ki-67 after fixation and permeabilization. Data were acquired on a Sony ID7000.

### ELISpot assays

IFN-γ ELISpot: Plates were coated with an anti-mouse IFN-γ capture antibody overnight. After blocking, splenocytes (5×10 cells/well) were added and stimulated with SARS-CoV-2 spike peptide pools (2 μg/mL) for 24 hours. Spots were developed using a biotinylated detection antibody, streptavidin-HRP, and an ACE substrate. Spots were counted using an automated ELISpot reader.

SARS-CoV-2-specific B cell ELISpot: Plates were coated with recombinant spike protein (10 μg/mL). After blocking, splenocytes (1×10 cells/well) were added and incubated for 24 hours. Antigen-specific IgG-secreting cells were detected using an anti-mouse IgG antibody, followed by an HRP-conjugated secondary antibody and ACE substrate.

### Secreted alkaline phosphatase (SEAP) assay

Serum samples were heat-inactivated at 56°C for 30 minutes. SEAP levels were measured using a colorimetric substrate assay according to the manufacturer’s instructions. Absorbance was measured at 405 nm, and concentrations were calculated from a recombinant SEAP standard curve.

### *In vivo* bioluminescence imaging

Mice were injected intramuscularly with 1×10¹ vp of Ad5 expressing firefly luciferase (Ad5-FLuc). At indicated time points post-injection (24–120 hours), mice were injected intraperitoneally with D-luciferin (150 mg/kg) and imaged under isoflurane anesthesia using an In Vivo Imaging System (AniView100 Pro, BLT). Bioluminescence signals were quantified as total flux (photons/second) within a defined region of interest.

### Detection of adenoviral genomes and transgene expression

DNA qPCR: Muscle tissue from the injection site was harvested at days 1, 7, and 14 post-Ad5-Spike injection. Genomic DNA was extracted, and adenoviral genome copies were quantified by qPCR targeting the hexon gene.

RNA qRT-PCR: Total RNA was isolated from snap-frozen muscle tissue using Trizol reagent. cDNA was synthesized, and the expression of the spike transgene mRNA was quantified by qPCR.

### Bulk RNA-sequencing and analysis

Draining lymph nodes were harvested from immunosuppressed mice three days post-immunization. Total RNA was extracted, and libraries were prepared from high-quality RNA (RIN ≥ 7). Sequencing was performed on an Illumina platform. Raw reads were quality-controlled and aligned to the mouse reference genome (*Mus musculus*, Ensembl v115) using Hisat2. Gene counts were generated with featureCounts. Differential expression analysis was performed with DESeq2 (v1.40.2) with default parameters. Genes with an adjusted P value (padj) < 0.05 and absolute log2 fold change > 1 were considered differentially expressed. Gene Set Enrichment Analysis (GSEA) and functional enrichment of co-expression clusters (identified by Mfuzz) were conducted using the clusterProfiler package and databases from MSigDB (GO, KEGG, Reactome).

### Statistical analyses

Statistical analyses were performed using GraphPad Prism 9.5.0 (GraphPad Software). Data are presented as median with interquartile range (IQR) and represent duplicate measurements. Comparisons between two groups were performed using two-sided Mann-Whitney U tests. A p-value less than 0.05 was considered statistically significant.

## DATA AVAILABILITY STATEMENT

All data needed to evaluate the conclusions in the paper are present in the manuscript and/or the Supplementary Materials. Any additional data is available on request from the corresponding author, Dr. Jingyou Yu at.

## ACKNOWLEDGMENTS

The authors thank members of the Yu and Zhu laboratories for their reagents, technical assistance, and insightful discussions. This study was funded by the National Key R&D Program of China (No. 2022YFC2304202 to Z.H.), Major Project of Guangzhou National Laboratory (GZNL2023A01009 and GZNL2023A01005 to J.Y.), the Major Research Plan of the National Natural Science Foundation of China (92469104 to J.Y.) and the National Natural Science Foundation of China (NSFC 82371831 to J.Y.).

## AUTHOR CONTRIBUTIONS

J.Y. and Z.Z. designed the study. J.Y., Z.Z., Y.B. and Z.H. supervised the whole study. S.L. led the immunologic, virologic, and animal work. W.Z. and J.F. led the clinic work under supervision of Z.Z. Bioinformatic analysis was led by C.X. The paper was written by J.Y., S.L., C.X. with the involvement of all coauthors.

